# Impact of postnatal depression on neonatal outcomes: an exploratory study in Kisumu County, western Kenya

**DOI:** 10.1101/2022.10.25.22281500

**Authors:** Catherine Gribbin, Florence Achieng, Alloys K’Oloo, Hellen C. Barsosio, Edith Kwobah, Simon Kariuki, Helen M Nabwera

## Abstract

**Background:** Previous Kenyan studies suggest postnatal depression (PND) may negatively impact infant growth. However these studies are limited to Nairobi and no research has explored the effects of PND in the neonatal period.

**Aim:** To explore the impact of PND on neonatal feeding practices, weight gain, illness episodes and identify key maternal caregiving challenges.

**Methods:** A mixed methods study of mothers and newborns <72 hours post-delivery from postnatal wards and clinics across 5 facilities in Kisumu County. At baseline, the Edinburgh Postnatal Depression Scale (EPDS) identified mothers with depressive features (EPDS ≥12) and infant feeding practices were assessed by questionnaire. 24 mothers were followed up 2 weeks later with a questionnaire and interview to further explore caregiving practices. Quantitative data was analysed using descriptive statistics. A thematic framework was used to identify and analyse emerging themes from the interviews.

**Results:** 56 (37%) out of 150 mother-infant pairs screened at baseline had an EPDS score ≥12. These mothers practiced exclusive breastfeeding less frequently (76.9% vs 90.9% p = 0.6) and a smaller proportion of their infants gained weight at 2 weeks (23.1% vs 36.4% p = 0.75). Key stressors were financial insecurity and lack of social support. Mothers described the benefits of social support on their mood and caregiving abilities.

**Conclusion:** Adverse growth and feeding outcomes are already apparent in the first 2 weeks of life among infants of mothers with features of PND. Early screening and intervention through community support structures could mitigate against the impact of PND on maternal mood and caregiving ability.

## Introduction

Postnatal depression (PND) is the most common complication of childbearing (1) and can seriously reduce a mother’s ability to care for herself and her infant (2). It is a significant contributor to maternal morbidity and mortality in low- and middle-income countries (LMICs) and is associated with adverse health and growth outcomes in infants (3-8). The prevalence of PND is twice as high in LMICs (20%) compared to high income countries (10%, HICs) (2). Whilst psychological interventions such as cognitive behavioural therapy have been shown to be effective in managing PND in HICs, these are rarely prioritised in LMICs due to the focus on addressing acute physical causes of maternal mortality in the postnatal period such as postpartum haemorrhage (9). Therefore, healthcare services in LMICs such as Kenya are ill-equipped to identify and manage mothers with PND.

In addition, the disability associated with PND is likely to impair maternal caregiving abilities for newborns at the most vulnerable time in their lives. Interventions which integrate both maternal and infant components are most successful at improving infant outcomes (9) thus it is important to explore both neonatal outcomes and maternal caregiving challenges associated with PND perspective before the relevant interventions can be developed.

Recent observational research from Kenya suggests the prevalence of PND may be higher among adolescent mothers (58%) (10) or those with acutely malnourished (64%) (4) and preterm infants (44%) (11, 12). However, none of the studies have explored how PND influences maternal infant care practices, particularly in the neonatal period where the infant is most vulnerable and essential feeding and care practices are established. Two weeks is a critical time for assessing infant progress post-discharge since low birth weight (LBW) infants are expected to have regained weight at this point which coincides with recommended postnatal follow-up visits and maternal well-being checks (13). Identifying the key caregiving challenges from the mother’s perspective is crucial in the design and implementation of targeted interventions. In addition, the majority of previous studies in Kenya were conducted in urban settings, whereas there is a need to explore the impact of PND in rural communities where mothers face different socio-economic pressures and healthcare quality and access is often poorer (14).

This study therefore aimed to explore how PND influences neonatal outcomes among mothers and their infants in urban and rural communities in Kisumu County of western Kenya.

## Methods

### Study design

This was a mixed methods study with two-stages of data collection based on the explanatory sequential design (15) **(Figure 1)**. At 0-72hrs post-delivery, mothers from postnatal wards and clinics were screened for PND using the EPDS and infant birthweight, health and feeding characteristics were determined with standardised questionnaires. Analysis of this data informed the purposive sampling of 28 mothers with and without PND who had characteristics of interest such as delayed breastfeeding initiation, LBW infants or those requiring admission to the neonatal unit. Four of these mothers were lost to follow-up. The remaining 24 mothers were followed-up in the community 2 weeks later with a second questionnaire assessing infant development and a semi-structured interview to further explore their caregiving experiences in the immediate postnatal period.

**Figure 1.**
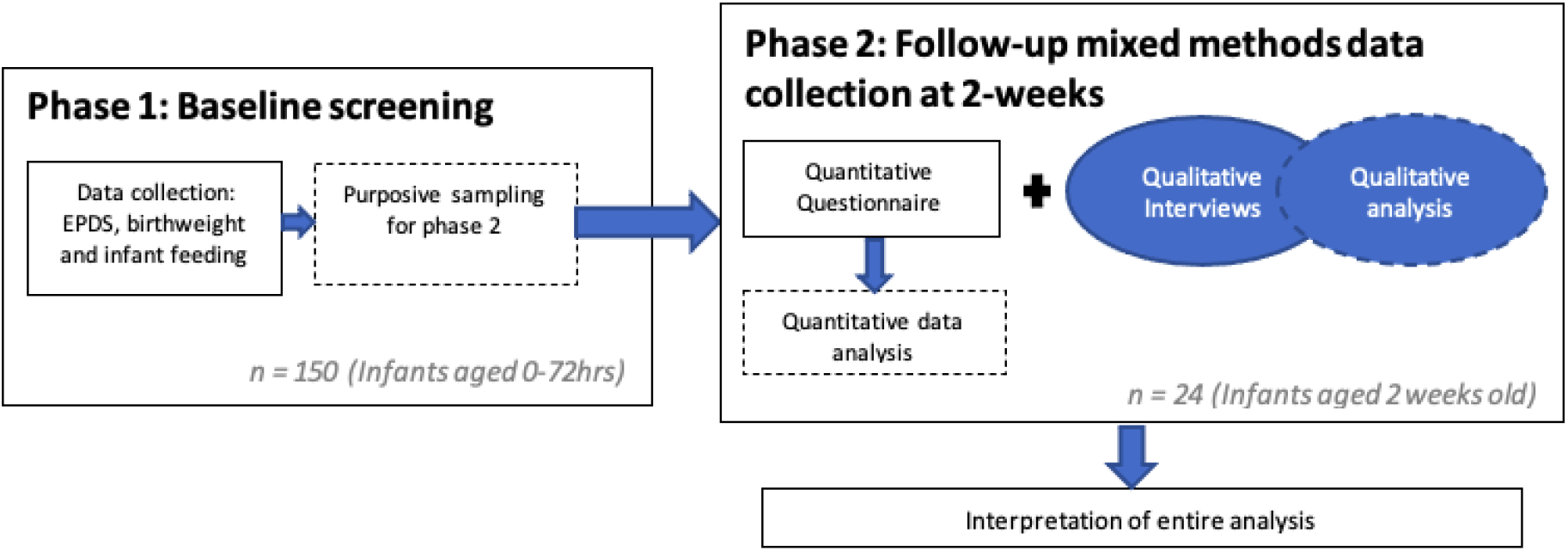
Study Design

**Figure 2.**
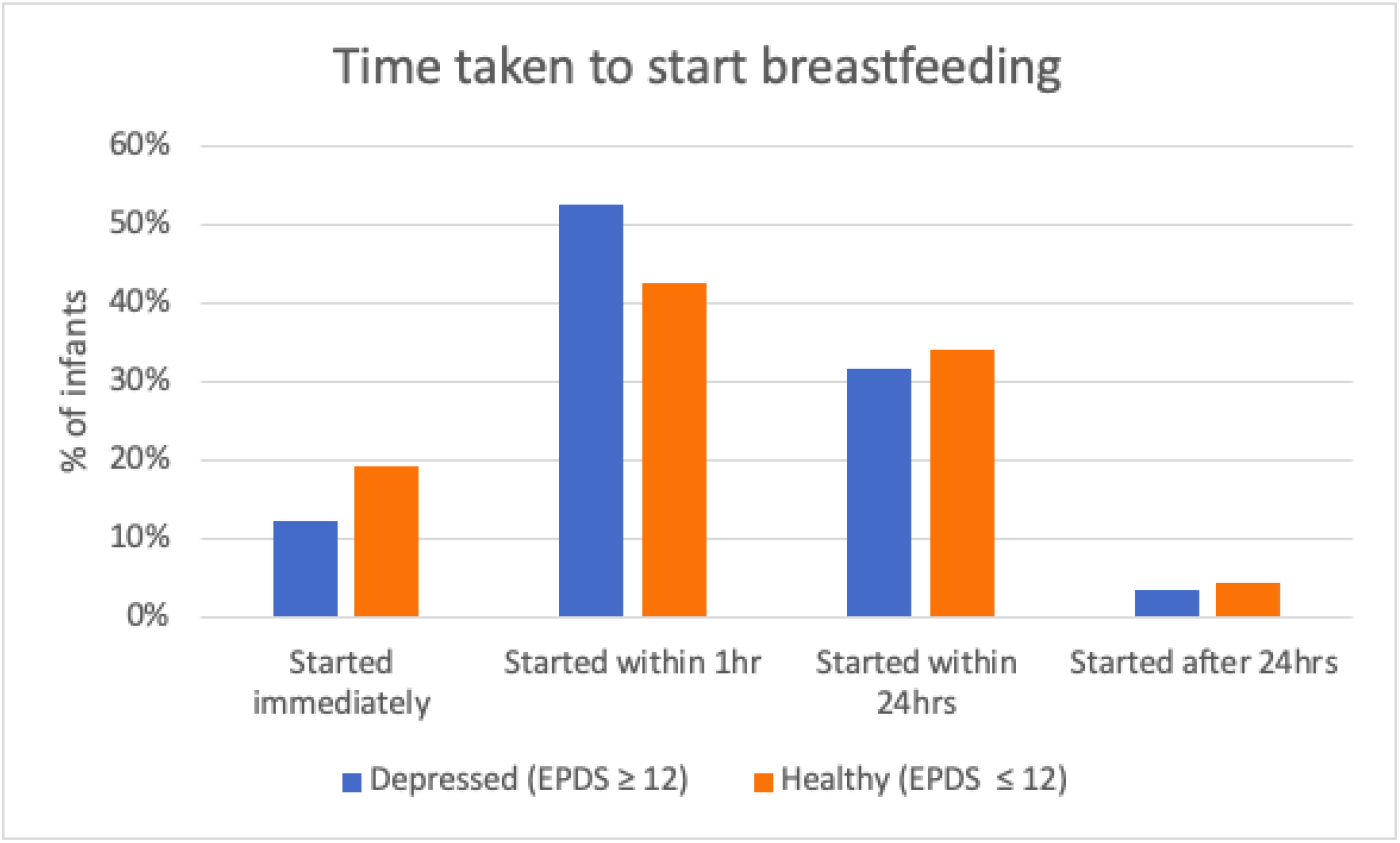
Time taken to start breastfeeding

### Study setting

The study was conducted in five health facilities across Kisumu County: Kisumu County Referral Hospital (n=52), Chulaimbo (n=19), Ahero (n=21), Rabuor (n=28) and Kombewa sub-County Hospitals (n=30). These facilities are embedded within an existing platform of maternal health collaborative research between KEMRI and LSTM. The lake region residents are predominantly of the Luo ethnic group, whose common livelihoods include agriculture, fishing, and microenterprise with both rural and urban communities (16). Over 60% of the households in Kisumu are estimated to be below the poverty line. Both infant and maternal mortality rates are significantly higher in Kisumu than Kenyan national averages (54 per 1,000 live births and 495 per 100,000 live births respectively) and Kisumu has the highest rate of teenage pregnancies in Kenya at 22% (17).

### Study population, sampling, and sample size

#### Baseline screening

In the first 2 weeks of data collection, we used convenience sampling (18) to recruit mothers from postnatal wards and clinics in the relevant health facilities, ensuring home deliveries were captured. All mothers with a live birth within 72 hours who gave informed consent were recruited by trained study staff. Mothers who had a still birth, refused follow-up or resided outside of Kisumu county were excluded. Study enrolment was voluntary, and participants were reimbursed for travel expenses according to Kenya Medical Research Institute-Centre for Global Health Research (KEMRI-CGHR) procedures. Our sample size of 150 mothe*rs* was based on the estimated prevalence of PND in Kenya of 18% (3) and the combined number of deliveries across study sites.

#### Follow-up data collection

##### Qualitative

Analysis of the screening data informed the purposive sample of 28 mothers for follow-up semi-structured interviews 2 weeks later. We included equal numbers of mothers with and without features of PND based on their EPDS scores and characteristics of interest including delayed breastfeeding initiation, non-exclusive breastfeeding, poor weight gain or infant illness. Mothers who scored ≤11 were considered ‘healthy’ for the purposes of this study. Including healthy mothers allowed us to identify similarities and differences in neonatal outcomes and maternal caregiving between the two groups whilst highlighting potential mitigating and exacerbating factors related to PND and its influence on caregiving. Sample size was based on achieving ‘data saturation’ where no new themes were emerging during the interviews (19). The sample comprised 13 mothers with features suggestive of PND (EPDS ≥12) and 11 healthy mothers (EPDS ≤11) as 4 mothers were lost to follow-up due to incomplete contact details.

##### Quantitative

Based on the sample size for follow-up qualitative data collection, we generated descriptive statistics to describe neonatal health, growth and feeding outcomes for these mothers and their infants at 2 weeks.

### Data collection

#### Baseline Screening

At baseline, the EPDS was used to screen 150 mothers over 2 weeks between May to June 2021, to identify those with features of PND (EPDS ≥12). This is a 10-item self-reporting questionnaire assessing recent emotional experiences using a numerical scale, that has been validated for use in Kenya and has been translated to the national language Kiswahili. For this study it was verbally translated and was read aloud to the mothers by a trained member of the study team in the local language, either Kiswahili or Dholuo. The target of 150 mothers was chosen for screening to identify a sufficient number with features of PND based on the expected prevalence (18%) (3). Mothers scoring ≥12 (total score 30) were classified as having symptoms suggestive of PND, and were referred to existing psychological services in their nearest County hospital. All mothers also completed a baseline questionnaire to assess feeding practices (including method, breastfeeding initiation, and complementary feeding), immunizations and special care required after birth such as resuscitation. Infant birth weight was recorded from hospital records and measured at baseline using calibrated scales available in the hospital facility.

#### Follow-up data collection

##### Quantitative

When infants were two weeks old, a second questionnaire was administered to 24 mothers at home during community follow-up visits. Here, basic maternal demographic data was collected and infant weight gain, feeding practices, illness episodes and maternal health seeking behaviour were assessed. Infant weight was measured during these visits to the nearest 10g by a trained member of the study team using calibrated digital (SECA 345) weighing scales.

##### Qualitative

Semi-structured interviews lasting approximately one hour were conducted by three research assistants (FA, AN and MAJ) who were all fluent in the local dialects-Dholuo and Kiswahili. Each mother was interviewed at home in the community by an individual interviewer, all of whom were female, with over 10 years’ experience in conducting qualitative interviews in maternal and child health research. All research assistants were trained by the study PI on data collection and ethical considerations. Topic guides used to structure the interviews comprised of open-ended questions addressing the following topics: feeding practices, maternal-infant interaction and attachment, infant illness episodes, support networks and maternal health. Interviewers documented field notes and recorded the interviews before transcribing them on the same day and translating them into English. Transcripts were reviewed by two interviewers to improve translation accuracy and were compared to the original audio recordings and revised appropriately following discussion where discrepancies arose. Regular participant checking helped to reduce the meaning lost through misinterpretation and enhance trustworthiness by prioritising the participants perspective.

All study tools were piloted before use amongst mothers in Kisumu County Referral Hospital to ensure the feasibility of data collection.

### Analysis

#### Statistical analysis

We used descriptive statistics to summarise the primary variables of interest including infant weight gain, feeding practices and illness episodes for mothers with features of PND and healthy mothers in our sample. Chi-square tests were applied to assess the similarity of these two groups.

#### Qualitative

We used the framework approach for qualitative data analysis to explore emerging themes from the interviews and investigate possible mechanisms of how PND impacts neonatal care and outcomes (20). Qualitative data analysis ran concurrently with data collection and was conducted by CG and FA. This involved familiarising ourselves with the data by re-reading the transcripts to identify recurring ideas, issues and inconsistencies; creating a coding framework using a mixture of deductive codes from our topic guide and inductive codes emerging from the data and then applying these codes to all transcripts in an iterative process using NVivo software (March 2020) (21). Similar codes were grouped, summarised, and compared using a ‘charting’ technique, retaining the original data and examining the similarities and differences across cases and themes. Peer review of transcripts and codes increased reliability and reduce inconsistencies. Relevant quotes are presented under key themes using matrices which ensure data is presented in its original form in an easily accessible format (22).

### Ethical statement

Ethical approval for the study was obtained from the Kenya Medical Research Institute’s Scientific and Ethics Review Unit and the Liverpool School of Tropical Medicine Research and Ethics Committee. Written informed consent was obtained from all participants prior to data collection. Due to high levels of illiteracy in the study communities, participants incapable of providing written informed consent were verbally guided through the study consent procedures and indicated their consent with a thumbprint.

## Results

### Baseline screening

Of the 150 mother-infant pairs recruited at baseline, 56 (37.3%; 95% confidence interval [CI]: 29.9 – 45.4) had an EPDS score ≥12 that was suggestive of PND. Out of 151 infants, there was one set of twins, 51% of infants were male (n=77), 98.7% were delivered in a health facility (n=149), 10.6% were admitted to the neonatal unit (n=16) and 4% infants were LBW (n=6).

The mean (SD) infant birth weight among the mothers with symptoms suggestive of PND was 3160g (474.35) vs 3258.87g (462.62) among healthy mothers. A similar proportion of infants required admission to the neonatal unit among mothers with features suggestive of PND vs healthy mothers [10.5% (n=6) vs 10.6% (n=10) respectively]. The number of infants immunised at birth was also similar [42.1% (n=24) and 42.6% (n=40) respectively].

Half of mothers with features of PND (53%) fed their infants within an hour of birth compared to healthy mothers 43% (p = 0.6) although healthy mothers were more likely to breastfeed their infants immediately after birth (19% vs 12% p= 0.6) (Error! Reference source not found..

At baseline, 94 (100%) healthy mothers reported that they practiced exclusive breastfeeding compared to 52 (93%) mothers with features suggestive of PND; 4 of whom reported feeding their infants both breastmilk and water (p = 0.6). These mothers used a cup to feed their infants expressed breastmilk and water rather than putting their baby on the breast as practiced by all healthy mothers. Three (23.1%) of the mothers with features of PND reported giving their infants additional fluids including saline and packet milk before breastfeeding whereas only 1 (9.1%) mother without features of PND gave her infant prelacteal fluids before breastmilk.

### Two-week follow-up data

#### Quantitative

*At two weeks, 24 (16%) mothers were then followed up: 13 with symptoms suggestive of PND versus 11 healthy*. Most mothers were married, and their mean age was 24 (SD 6.7) years. Trading was the most common source of income amongst participants although a larger proportion of ‘healthy mothers’ reported that their monthly income was guaranteed compared to mothers with depressive symptoms (63.6% (n = 7) vs 46.2% (n=6) p = 0.45). Similarly, 36.4% (n= 4) of ‘healthy’ mothers had attained post-secondary school education levels compared to only 7.7% (n = 1) of mothers with symptoms suggestive of PND (p = 0.5) **(Table 1)**.

**Table 1.** Participant characteristics

| *Table 1: Participant characteristics* |  |  |  |
| --- | --- | --- | --- |
| Variable | Depressed (EPDS $\geq 12$) | Healthy (EPDS $\leq 11$) | $\chi^2$ p-value |
| Marital Status | n (%) | n (%) |  |
| Never married | 3 (23.1) | 2 (18.2) |  |
| Currently married | 10 (76.9) | 9 (81.8) | 1.00 |
| Maternal age (years) |  |  |  |
| < 20 | 4 (30.8) | 2 (18.2) |  |
| 21 – 30 | 9 (69.2) | 9 (81.8) | 0.65 |
| Source of income |  |  |  |
| Farming | 1 (7.7) | 0 (0.0) |  |
| Trading | 8 (61.5) | 4 (36.4) |  |
| Salary | 2 (15.4) | 3 (27.3) |  |
| Remuneration from family member | 1 (7.7) | 2 (18.2) |  |
| Other (Odd jobs) | 1 (7.7) | 2 (18.2) | 0.67 |
| **Income guaranteed every month** |  |  |  |
| Yes | 6 (46.2) | 7 (63.6) | 0.45 |
| **Education level** |  |  |  |
| No schooling or did not complete primary school | 5 (38.5) | 3 (27.2) |  |
| Primary school completed or secondary school started but not completed | 5 (38.5) | 3 (27.2) |  |
| Secondary school completed | 2 (15.4) | 1 (9.1) |  |
| Tertiary or post-secondary school education | 1 (7.7) | 4 (36.4) | 0.50 |
| **Infant gender** |  |  |  |
| Male | 7 (53.8) | 7 (63.6) |  |
| Female | 6 (46.2) | 4 (36.4) | 0.70 |

#### Infant feeding practices

At 2 weeks, all mothers reported that they practiced exclusive breastfeeding (EBF) except one mother with features suggestive of PND whose infant was mixed fed on breastmilk and formula milk **(Table 2)**. The majority of mothers in both groups fed their infants over 8 times a day although interestingly, 27.3% healthy mothers (n= 3) reported feeding their infants only 1 to 4 times a day whilst 100% mothers with features of depression fed their infants at least 5 times a day.

**Table 2.**
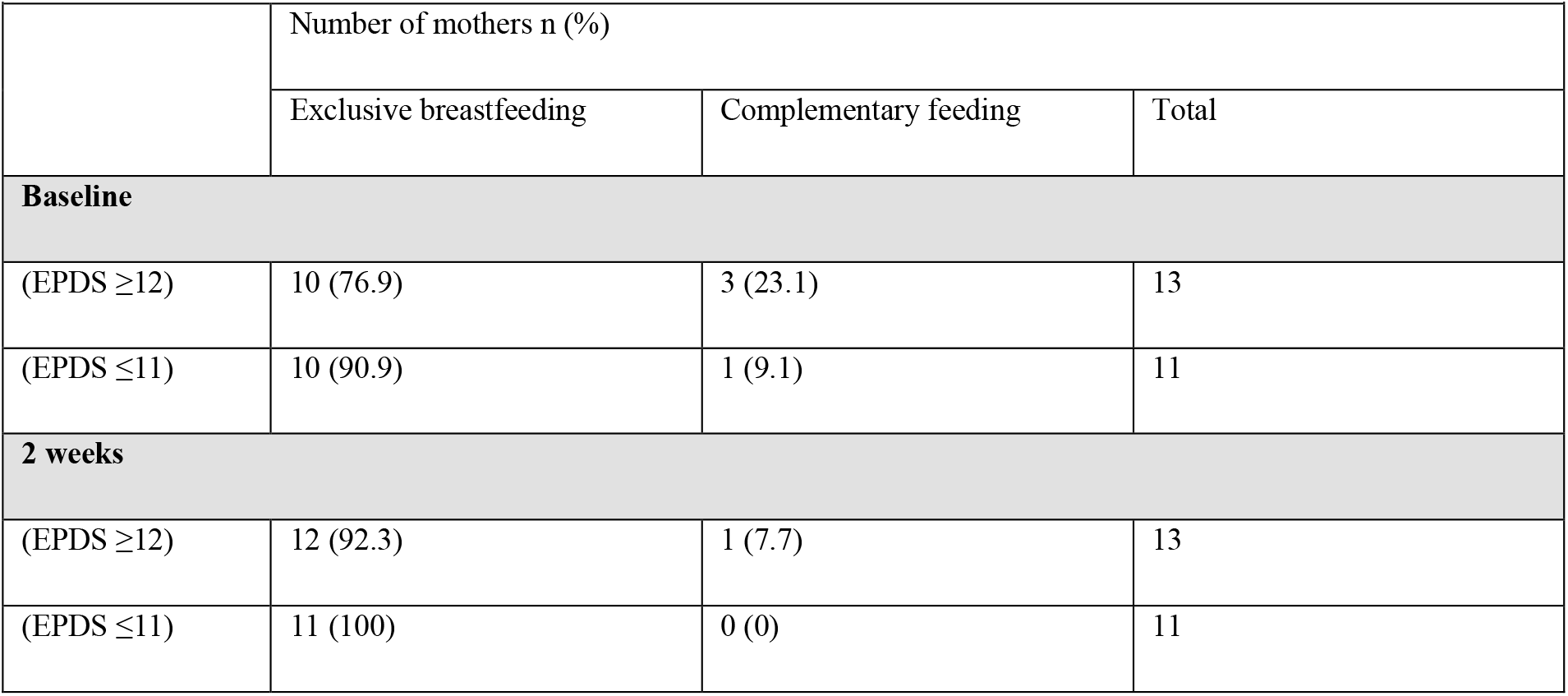
Feeding at baseline and 2 weeks

| *Table 2: Feeding at baseline and 2 weeks* |  |  |  |
| --- | --- | --- | --- |
|  | Number of mothers n (%) |  |  |
|  | Exclusive breastfeeding | Complementary feeding | Total |
| **Baseline** |  |  |  |
| (EPDS $\geq 12$) | 10 (76.9) | 3 (23.1) | 13 |
| (EPDS $\leq 11$) | 10 (90.9) | 1 (9.1) | 11 |
| **2 weeks** |  |  |  |
| (EPDS $\geq 12$) | 12 (92.3) | 1 (7.7) | 13 |
| (EPDS $\leq 11$) | 11 (100) | 0 (0) | 11 |

#### Infant weight gain

There was no difference in the mean weight at birth vs 2 weeks among infants of mothers with features suggestive of PND (p= 0.75). However, over the same interval, in infants of “healthy” mothers the mean weight increased by 66.1g (SD 206.8) (**Figure 3:** Infant weight from birth to two weeks

**Figure 3.**
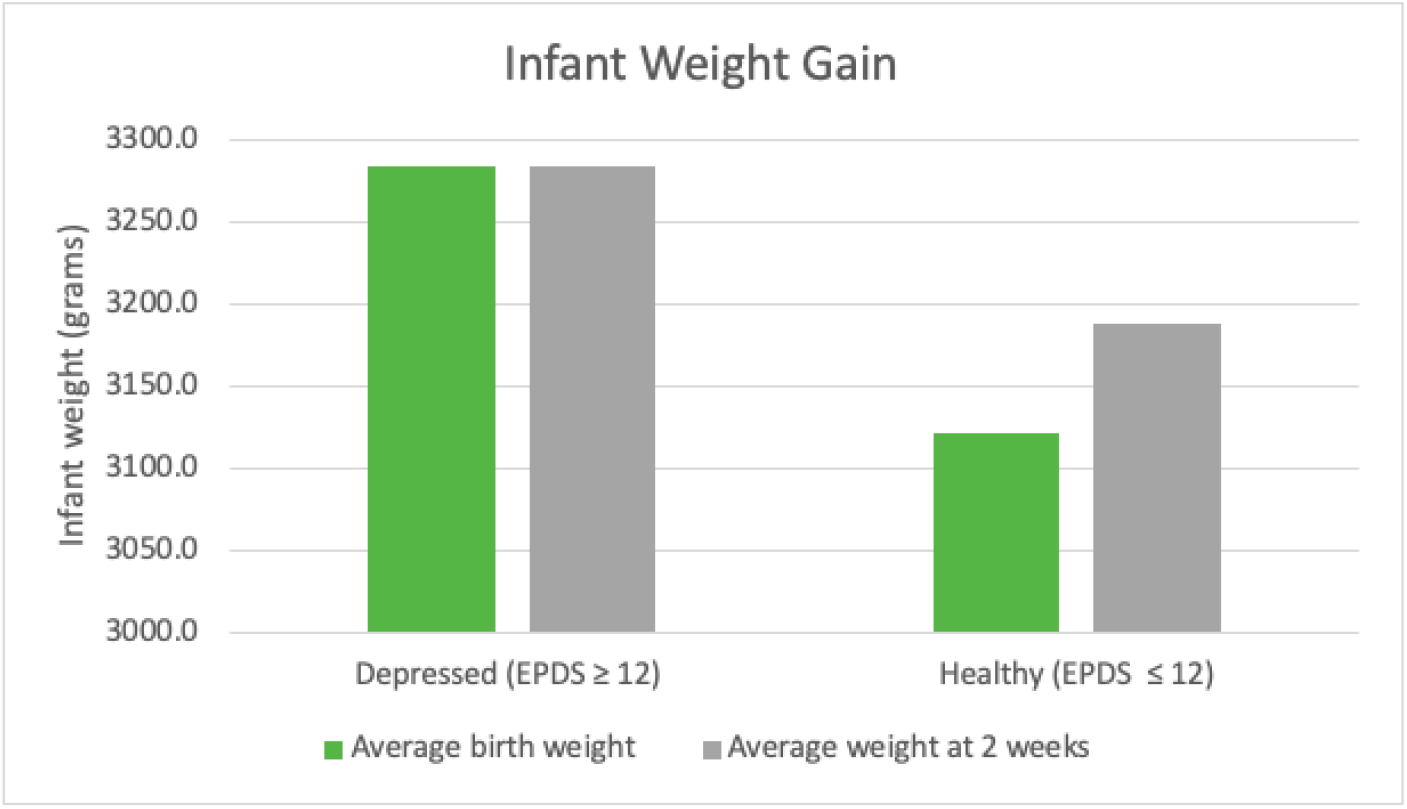
Infant weight from birth to two weeks

#### Infant health at 2 weeks

At 2 weeks of life, 2 (15.4%) mothers with features of PND reported an episode of infant illness vs 1 healthy mother (9.1% p=0.48) and all 3 mothers attended a health facility. In total, 5 (38.5%) mothers with features suggestive of PND vs 4 (36.4%) healthy mothers visited a health facility although the predominant purpose of these visits was for infant immunizations rather than seeking care for health problems **(Table 3)**.

**Table 3.** Number of infant illness episodes and health facility visits

| *Table 3: Number of infant illness episodes and health facility visits* |  |  |
| --- | --- | --- |
| Variable | Depressed (EPDS ≥ 12) | Non-Depressed (EPDS $<$ 12) |
| **Number of episodes baby unwell** | **n (%)** | **n (%)** |
| None | 11 (84.6) | 10 (90.9) |
| Once | 2 (15.4) | 0 (0) |
| Twice or more | 0 (0) | 1 (9.1) |
| **Visits to health facility** | **n (%)** | **n (%)** |
| None | 8 (61.5) | 7 (63.6) |
| One to two | 5 (38.5) | 4 (36.4) |

#### Qualitative

All mothers within our study were exposed to health or socio-economic adversities in the intrapartum and postnatal period; be it traumatic delivery, financial constraints, or relationship difficulties. However, it remains unclear why some of these mothers developed features suggestive of PND in the immediate postnatal period whilst others did not. Interestingly, rather than depression or low mood, the mothers we interviewed were more likely to mention ‘overthinking’, often linked to the aforementioned adverse circumstances.

*“I just think that its overthinking that is what makes us tired…I have too much to think about…Thinking about how you want to look for money to help yourself that is the main thing that is making me think a lot” (RH-MO25)*. We developed the Conceptual Framework in **Figure 4** to explore the how maternal PND could influence infant care practices and outcomes.

**Figure 4.**
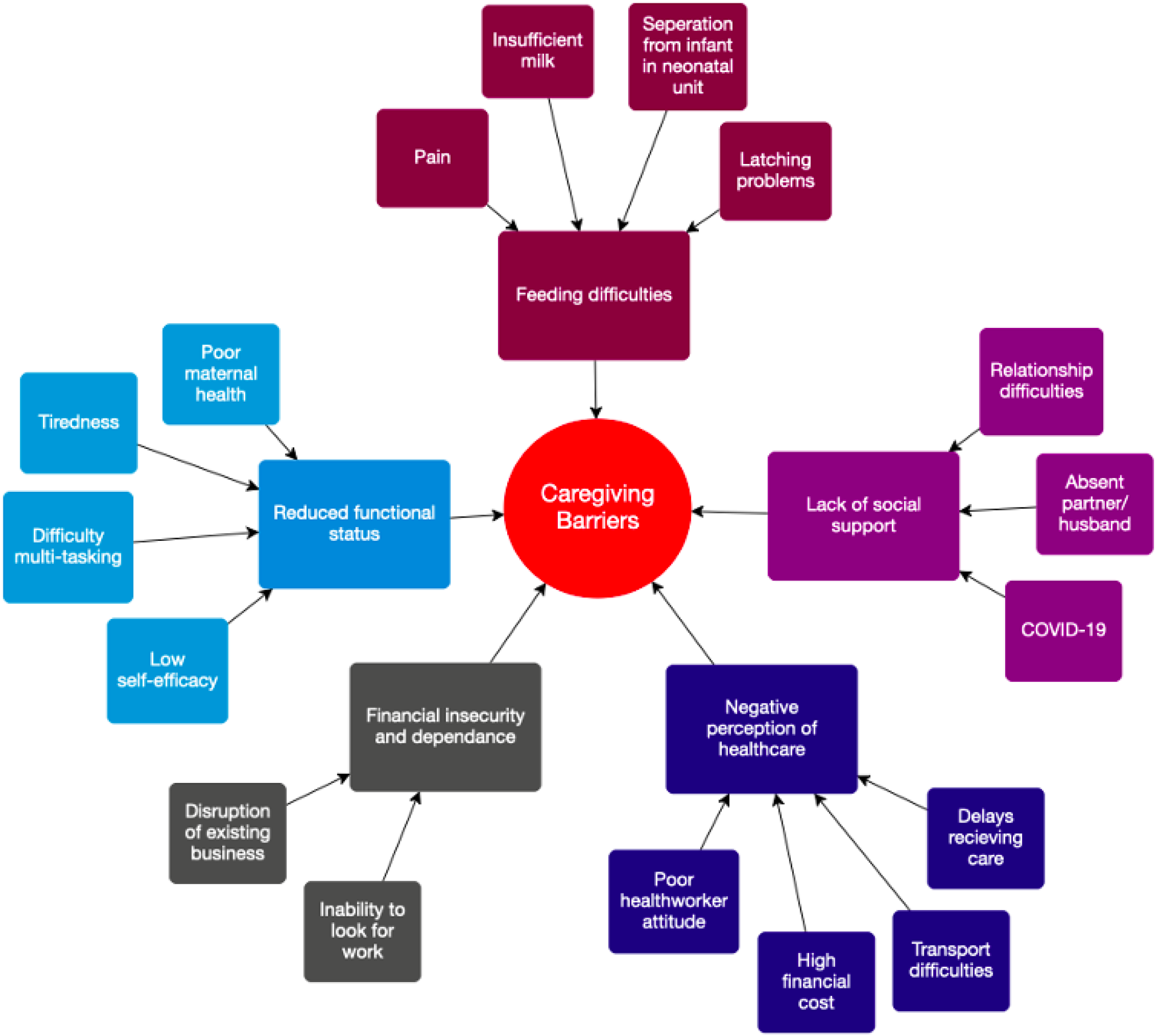
Conceptual framework of caregiving barriers

#### The impact of PND on feeding practices

All mothers demonstrated responsive feeding practices by describing an awareness of infant hunger and satiety cues such as their infant ‘wanting to suck the fingers’ or ‘pulling out the tongue’. Mothers placed huge emphasis on infant satiety given its nutritional benefits. Both mothers with features suggestive of PND and healthy mothers described similar feeding difficulties including problems with infants latching, pain due to breast engorgement, and the practical and emotional challenges associated with feeding an infant requiring care in a neonatal unit (**Figure 4**, **Table 4**Table 4**: Q1-3**). Insufficient milk was another emerging sub-theme, particularly amongst mothers with features suggestive of PND and was also given as a justification for introducing complementary feeds Mothers disputed whether poor breastmilk production was due to the infant taking too much milk or a result of poor maternal diet **(Table 4: Q5-6)**.

**Table 4.** Feeding challenges

| *Table 4: Feeding challenges* |  |  |
| --- | --- | --- |
| Sub-themes |  |  |
| Latching problems | Q1 | *Whenever am breastfeeding her she feels like she wants to hold but she cannot hold it with the mouth because it is slippery...that is something I find hard (CH-MO14)* |
| Pain | Q2 | *"The only problem I have is my painful breast, it worries me whenever I want to breastfeed, and it was very painful that I had to shed tears while breastfeeding but I just persevered" (RH-MO25)* |
| Separation from infant in neonatal unit | Q3 | *"Walking up and down is not easy because I had to go upstairs to breastfeed my baby in the nursery and I had just had an operation" (KC-MO26)* |
|  | Q4 | *The challenge I had was going to the nursery to breastfeed my child and at the same time I had a lot of pains (AC-MO07)* |
| Insufficient milk | Q5 | *"When breastfeeding him I realized that the milk was not enough and that is the reason why I introduced milk" (RH-MO20)* |
|  | Q6 | *"with breastfeeding you need to eat well to get enough milk to feed the baby but sometimes I do not get enough food to eat. Like at times there is no proper food to eat and at times you just cook porridge and drink which is not enough" (RH-MO04)* |

#### The impact of PND on infant health and maternal health-seeking behaviour

Both groups of mothers described their infants experiencing ‘stomach-ache’, often leading to disrupted sleep for both mother and infant. Mothers also raised the high financial cost and practical challenges associated with travelling long distances to the nearest health facility (**Figure 4**, **Table 5 Q7, Q8, Q9**), often to be met with long waiting times and supply shortages resulting in delays receiving care **(Table 5:** Q10). When mothers did access healthcare, they reported a lack of information from healthcare professionals regarding their own treatment for example during delivery, or the healthcare needs of their child, which contributed to severe maternal worry and distress **(Table 5: Q11)**. One mother with depressive features described the nurses shouting and harassing mothers on the ward whilst another expressed that she could not go back to hospital following the maltreatment she had received. **(Table 5: Q11, Q12)**. Both participants highlighted the anxiety encountered by mothers experiencing childbirth alone **(Table 5: Q13)**.

**Table 5.** Health-seeking challenges

| *Table 5: Health-seeking challenges* |  |  |
| --- | --- | --- |
| Sub-themes |  |  |
| Financial cost | Q7 | *“...at times you want to go to the hospital, but you don’t have the money, so I walk” (AC-MO02)* |
|  | Q8 | *“...when I got there, they told me that they did not have the equipment’s to help me deliver that I should go and buy” (RH-MO25)* |
| Transport difficulties | Q9 | *“I did not go to the hospital because my husband was very busy, and he has to take me to the hospital. I am also not able to walk by myself because of my disability” (RH-MO20)* |
| Delays receiving care | Q10 | *“sometimes they don’t help the baby can end up dying when they delay with the treatment... it can even take three hours before you are attended to” (KC-MO04)* |
| Poor health-worker attitude | Q11 | *“The doctors did not attend to me well, and they made me feel so bad... she just took the baby and rushed with her to the nursery...I asked if the baby was going to be okay, but they just looked at me but did not answer” (KC-MO07)* |
|  | Q12 | *“they should at least tell us what to do instead of them rushing us we don’t need those nurses/trainees, we need those with experience, because they shout at us and harass us, it makes us depressed, like with my case the way the sisters handled me, they had an attitude towards me, but to men, they are just okay” (CH-MO14)* |
|  | Q13 | *“there was a scenario where young girl came with the mother, she was only 13 years, the mother was not allowed to the delivery room, so they shouted at her until the mother came to ask what was going on, the nurses just looked at her, so she told the nurse that that was not the only hospital she can take her elsewhere, that is when they changed, this acts makes us depressed so they should allow any female person to come with us in the delivery room” (CH-MO14)* |

#### The impact of PND on mother-infant interaction

Both groups of mothers recognised the importance of interacting with their infants and noted the positive effect of this on their mood. In general, this involved a combination of talking to, singing to, and touching their infant (**Figure 4**,**Table 6: Q14, Q15)**. However, some mothers expressed their frustration at the lack of infant response. This was particularly noticeable amongst healthy mothers, many of whom believed that their infants were too young to hear anything **(Table 6: Q16, Q17, Q18**). No participants reported they had been separated from their infants in the first two weeks, although one mother with depressive features explained the difficulty of ‘finding a good time’ to spend with her infant, given her burden of household tasks and other caregiving duties **(Table 6: Q19)**. Nevertheless, mothers prioritised infant needs; perhaps most evident in their response to infant crying where mothers described the urgency in which they would comfort their child by carrying or breastfeeding them **(Table 6: Q20, Q21)**.

**Table 6.**
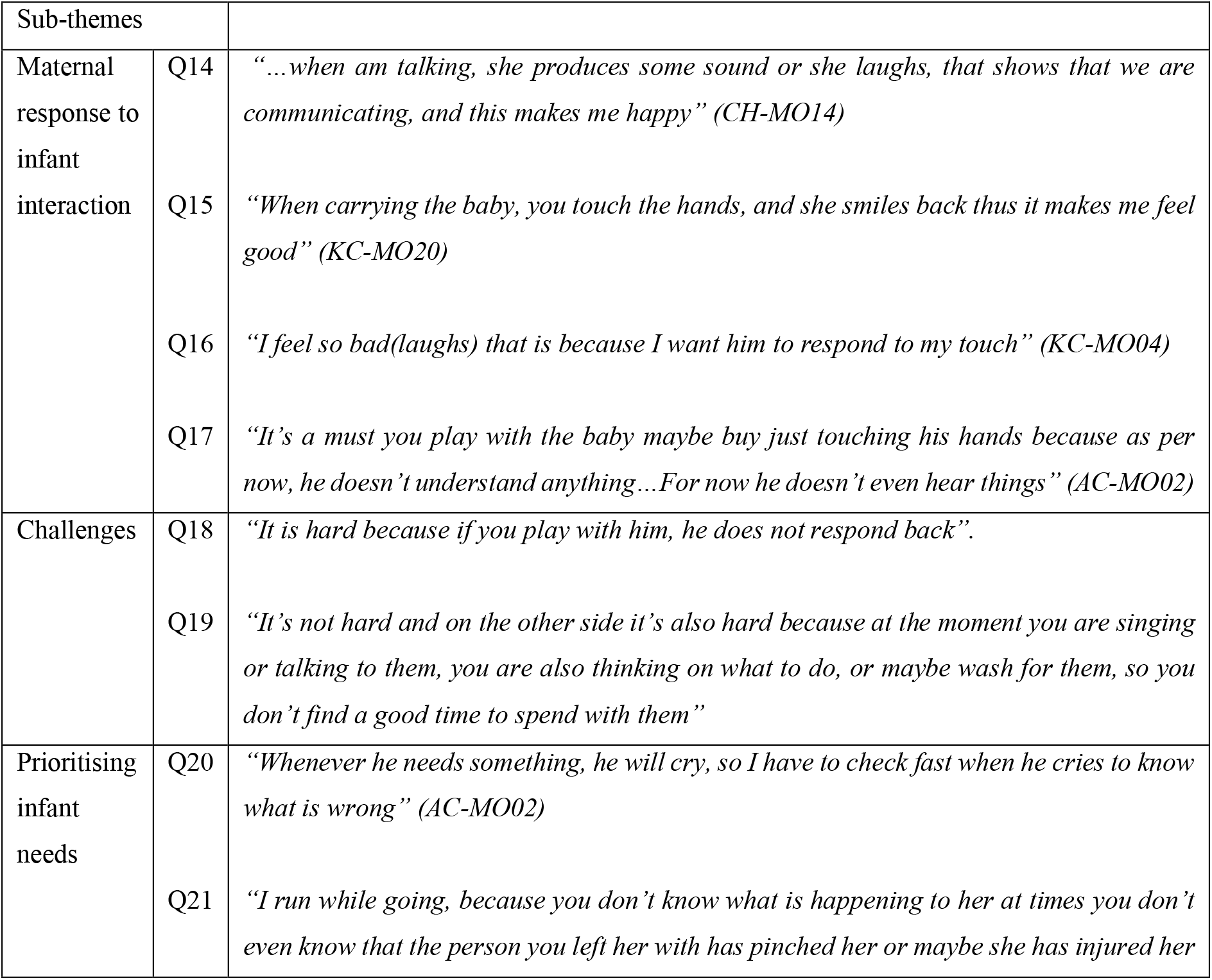
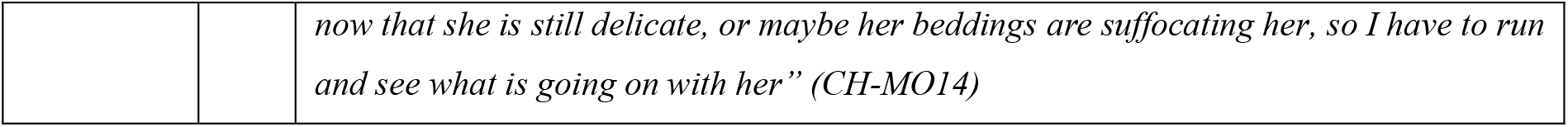
Mother infant interaction

#### The impact of PND on maternal self-efficacy and functional ability

When asked about conducting every day household tasks, all mothers described reducing their workload **(Table 7: Q22, Q23**). Despite this, mothers with features suggestive of depression were more likely to express that they felt unable to care for themselves and their infants compared to healthy mothers often because of poor social support or ill health **(Table 7: Q24)**. Interestingly, some mothers with stronger support networks were advised by relatives not to complete strenuous household jobs, suggesting that in some cases this lack of self-efficacy may be re-enforced by social support systems. Mothers in both groups also linked their lack of financial independence to poor self-efficacy **(Table 7: Q25, Q26)**.

**Table 7.** Maternal functional status

| *Table 7: Maternal functional status* |  |  |
| --- | --- | --- |
| Sub-themes |  |  |
| Reduced workload | Q22 | *“I just manage by doing my daily chores in bits and when tired I rest and start doing another thing again” (KC-MO04)* |
|  | Q23 | *“I just do light things even when washing I just do light clothes” (KC-MO24)* |
| Poor maternal health | Q24 | *“At times you find that you want to do some of the house chores, but you can’t because at that time she might be crying and maybe I was cooking, so I find this very hard because I can’t multitask...at the moment I can’t find a way of helping myself because I can’t go to work because of my health, I can’t stand or along time too with this I can’t help take care of myself... Now, I can only wash while seated I cannot bend yet, still I can’t cook; I can’t mop because when I do them, I feel so much pain on my thighs” (CH-MO14)* |
| Lack of financial dependence | Q25 | *“Apart from that is that you are just sited at home your ways are blocked, there is nothing you can do to sustain yourself because you can’t go with her anywhere, so you find that your business is at stand still, those are the difficult things I see” (KC-MO24)* |
|  | Q26 | *“Now that am not going to work it’s hard because the baby needs feeding, clothes to wear and soap to wash his clothes...I need help because right now there is nowhere going, but before I could go to the market to sell vegetables and from there, I could find how to help myself” (AC-MO02)* |

#### Social support

In general, husbands and partners were absent for most of the day although they had an important role in providing financially **(Table 8**Table 8**: Q27, Q28)**. Thus, unless mothers received support from neighbours, friends or relatives, they were alone caring for their infant and completing household tasks which mothers expressed could be difficult. Whilst some mothers received practical support including help with ‘cooking and cleaning’, giving them the opportunity to spend time with their infants, other mothers received psychological and emotional support, often in the form of advice **(Table 8: Q29, Q30, Q31, Q32)**. Mothers mentioned the positive impact of social support on their mood and implied that good social support prevented them from feeling low **(Table 8: Q33, Q34)**. One mother commented on how having a network of other new mothers allowed her to normalise and therefore cope with some of the challenges she was facing caring for a newborn (**Table 8: Q35)**. Despite these benefits, anxiety surrounding COVID-19 meant some mothers limited their support networks to prevent virus transmission to their infants **(Table 8: Q36, Q37)**.

**Table 8.**
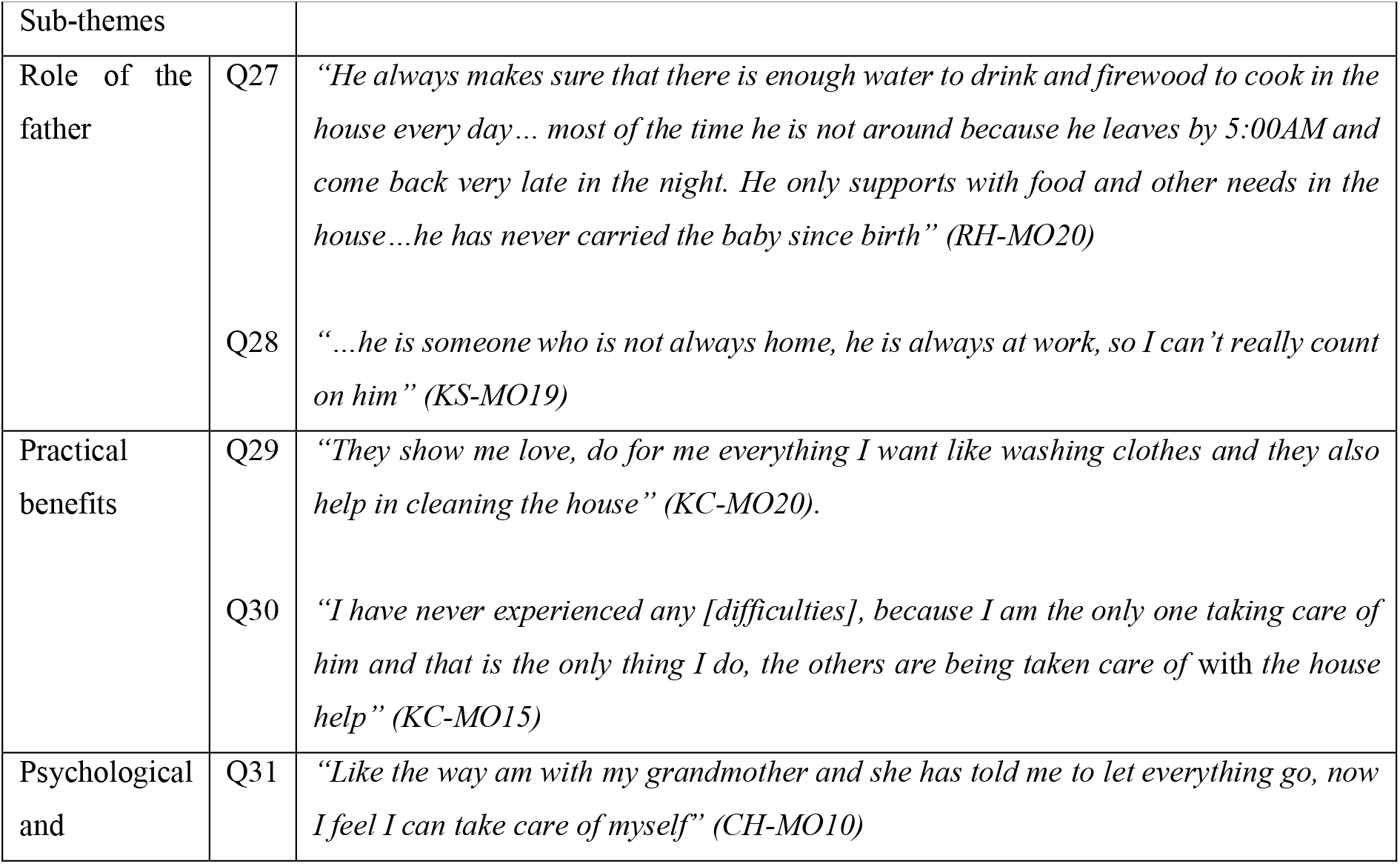
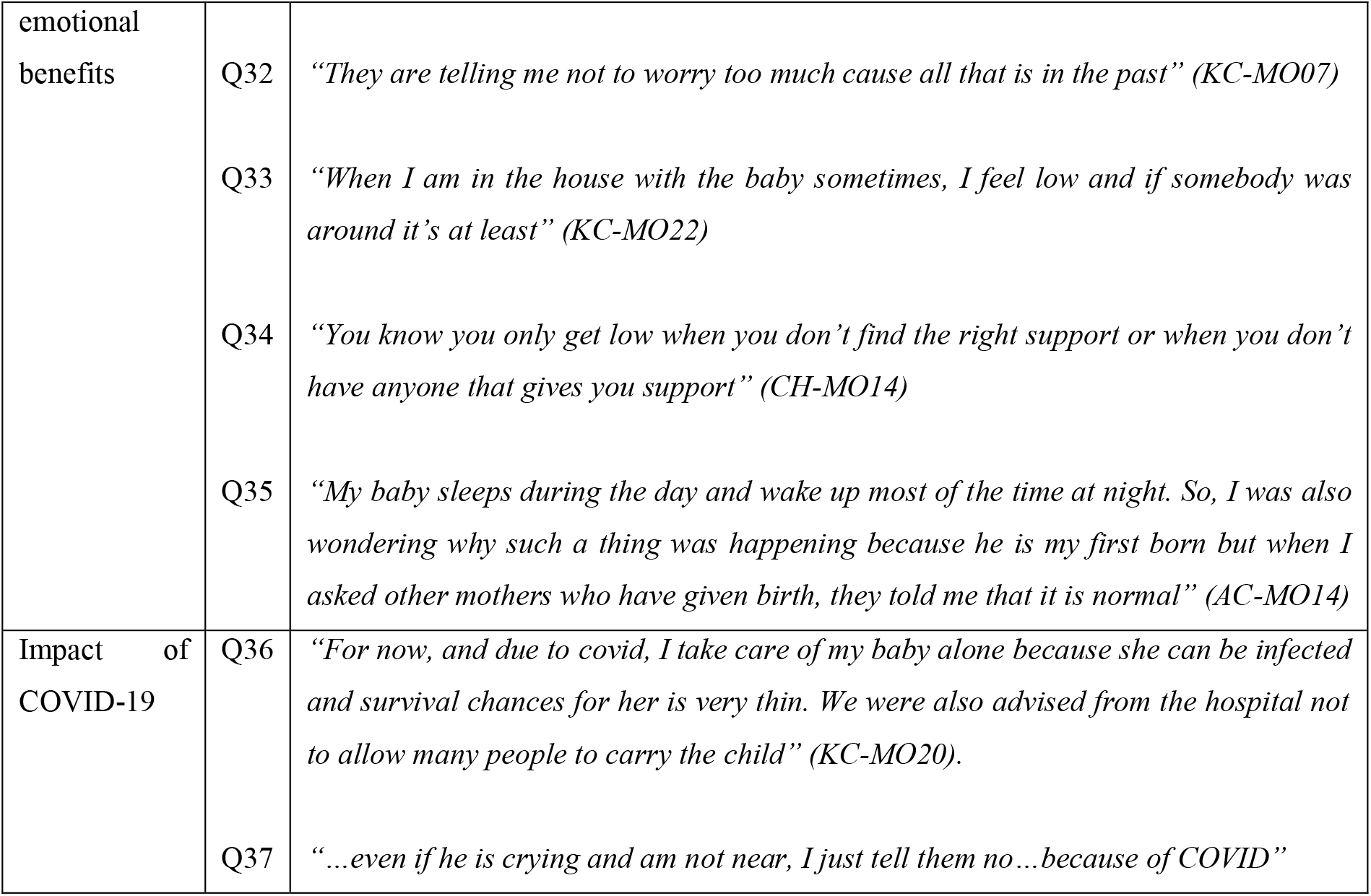
Social support

## Discussion

### Summary of key findings

Although not statistically significant, our findings suggest that mothers with features of PND practised EBF less frequently in the first 2 weeks of life and their infants’ weight gain was inadequate. Key caregiving challenges for mothers included feeding difficulties and reduced functional status due to poor physical health, psychological stress due to poor health care experiences, financial insecurity and lack of social support. Mothers described how practical and emotional support from partners, friends and relatives in the immediate postnatal period mitigated these challenges. There were no obvious disparities between the two groups in terms of mother-infant interaction, infant illness episodes and maternal health-seeking although many mothers reported negative and traumatic experiences surrounding childbirth.

### Impact of PND on breastfeeding and growth

Our findings suggest that the well-documented association between PND and poor infant growth is already apparent in the first 2 weeks of life, highlighting the need for early infant breastfeeding interventions. A potential explanation for poorer weight gain amongst these infants could be maternal undernutrition which has been linked to both PND and impaired foetal and early infant growth in the postnatal period (23). Whilst we did not directly assess maternal nutrition, multiple mothers in our study described insufficient breastmilk production which they attributed to poor diet. This was the main reason for non-EBF was more common amongst mothers with features of PND, consistent with previous research in Kenya (24). Furthermore, mothers with depressive characteristics fed their infants more frequently than healthy mothers which could indicate poor infant satiety. Alternatively, mothers with PND are more likely to have preterm or small for gestational age infants (25) which may explain the poor weight gain amongst these infants, although this would only be possible to ascertain using a longitudinal study.

Whilst research in HICs suggests that depressed mothers may encounter problems with breastmilk production due to reduced oxytocin and prolactin production (26), all breastfeeding measures in our study were self-reported, hence it is difficult to establish whether there was an objective difference in breastmilk production between the two groups or if this was biased by exaggerated pessimistic concern previously noted in depressed mothers (27). Instead, these beliefs about insufficient breastmilk may be due to low maternal self-efficacy which is more common amongst mothers with PND (28). Rahman et al. found no difference in breastmilk production between depressed and healthy mothers in rural Pakistan at 4 months (29), even though depressed mothers were more likely to report insufficient milk. This would fit with our qualitative findings which suggest that mothers with depressive features have less confidence in their ability to care for themselves and their infants despite describing similar degrees of functional disability as healthy mothers. Previous research in Iran has linked PND to reduced functional status and poor maternal self-efficacy(30) and breastfeeding self-efficacy training has been shown to significantly improve depression outcomes amongst these mothers (31).

Most mothers in both groups began feeding their infants within an hour of birth which is earlier than previous research suggests. A large observational study in India found a lower odds of EBF in mothers with PND in the first 48 hours after birth (32) which can lead to reduced breastfeeding success in the postnatal period (33). In contrast, a greater proportion of mothers with PND in our study were practicing EBF at 2 weeks chronological age compared to at 72 hours although the reason for this remains unclear. Interestingly, all mothers who transitioned from complementary feeding to EBF attended a health facility in the first 2 weeks after delivery, hence these mothers may have received additional breastfeeding support and advice from trained health professionals. Research in Nigeria implies that early routine immunisation visits are often the only contact a mother will have with a health professional post-delivery and this may be similar in rural western Kenya where postnatal service uptake is poor (12). Thus, early immunisation visits could provide a key opportunity for reaching mothers and their newborns with breastfeeding interventions, particularly since our study shows both mothers with and without depression were likely to attend these clinics.

### Maternal well-being

Financial insecurity leading to economic stress, relationship difficulties and a lack of social support were the prominent factors mothers associated with ‘worry’ or ‘over-thinking’. Therefore, whilst breastfeeding and infant growth interventions are likely to have a positive impact on maternal mood (9, 34) interventions that aim to improve social support and financial security will be important to fully address the sources of maternal anxiety and stress in the postnatal period. Although it is difficult to minimise the adverse events encountered by these mothers in challenging social and financial circumstances, it may be possible to build their resilience to cope with such events namely through increasing maternal self-efficacy and strengthening maternal support networks. Likewise, mothers described intense labour pain, feelings of powerlessness, negative health worker interactions and being given a lack of information at the point of delivery; all of which are potentially modifiable factors contributing to traumatic childbirth experience (35), associated with the development of PND (36, 37).

### Impact of social support

Our findings suggest that social support had a positive effect on maternal mood and self-efficacy consistent with previous research (38). Practical assistance including help with housework and financial support allowed mothers to focus their effects primarily on infant caregiving and enabled mothers to rest and recover back to full health following delivery, which was considered important for good functional ability. In contrast, mothers who lacked practical support were worried about economic difficulties and food insecurity, an important and potentially modifiable risk factor for depression (39, 40). A study of South African mothers found that food insufficiency was more likely to compromise the mental health mothers who lacked social support and practical support was more effective than emotional support at mitigating these adverse effects on mental health (39). Nevertheless, mothers in our study described the benefits of receiving emotional support from relatives which allowed them to reframe negative thinking patterns, normalise caregiving challenges and boost self-esteem which is consistent with previous LMIC research (40, 41).

In our recent research exploring the feasibility of mother-to-mother peer support for the post discharge care of LBW infants, we found that this strategy promoted resilience among mothers of LBW infants, improved knowledge and practice of breastfeeding and other caregiving practices, and enhanced family relationships(42). Similarly a recent meta-analysis suggested community based peer-support such as one-to-one counselling or group meetings can increase breastfeeding initiation within the first hour and extend the duration of EBF as well as reducing prelacteal feeding amongst new mothers in LMICs (43).

### Limitations

The cross-sectional study design hinders our ability to understand the causal mechanisms behind the impact of PND on infant growth, breastfeeding and maternal caregiving. Longitudinal data mapping out maternal experiences across the entire perinatal period would be useful to further understand why some mothers develop PND and others do not as well as enabling us to truly isolate the impact of PND on maternal caregiving in the postnatal period by identifying antenatal confounding factors. Whilst this study provides useful exploratory data, the small convenience sample used prevents us from generalising our findings and comparisons between the two groups of mothers are not conclusive due to small sample size.

### Conclusion

Our findings suggest that mothers in western Kenya with features of PND in the immediate postnatal period may practise EBF and appropriate infant caregiving less frequently and lack social support. Early screening and intervention for PND could prevent maternal and infant adverse outcomes which already exist in the early postnatal period. In addition, health workers should receive sufficient training to enhance their interactions with mothers during pregnancy, delivery and postnatal periods. Finally, strengthening community-based social support networks could build maternal resilience and autonomy to enhance their psychosocial well-being with ultimate benefits for them, their infants and families.

## Data Availability

All data produced in the present study are available upon reasonable request to the authors

## Acknowledgements

We would like to thank all the mothers who participated in this study and health care workers who supported us with recruitment and follow-up of the mothers. Our gratitude goes to Dr Sarah White for her statistical input to the study design.

## References

1. Sit DKY, Wisner KL. Identification of postpartum depression. Clin Obstet Gynecol. 2009;52(3):456–68.

2. Woody CA, Ferrari AJ, Siskind DJ, Whiteford HA, Harris MG. A systematic review and meta-regression of the prevalence and incidence of perinatal depression. J Affect Disord. 2017;219:86–92.

3. Ongeri L, Wanga V, Otieno P, Mbui J, Juma E, Stoep AV, et al. Demographic, psychosocial and clinical factors associated with postpartum depression in Kenyan women. BMC Psychiatry. 2018;18(1):318.

4. Haithar S, Kuria MW, Sheikh A, Kumar M, Vander Stoep A. Maternal depression and child severe acute malnutrition: a case-control study from Kenya. BMC Pediatrics. 2018;18(1):289.

5. Adewuya AO, Ola BO, Aloba OO, Mapayi BM, Okeniyi JAO. Impact of postnatal depression on infants’ growth in Nigeria. Journal of Affective Disorders. 2008;108(1/2):191–3.

6. MacGinty R, Lesosky M, Barnett W, Nduru PM, Vanker A, Stein DJ, et al. Maternal psychosocial risk factors and lower respiratory tract infection (LRTI) during infancy in a South African birth cohort. PLoS One. 2019;14(12):e0226144.

7. Bennett IM, Schott W, Krutikova S, Behrman JR. Maternal mental health, and child growth and development, in four low-income and middle-income countries. Journal of Epidemiology and Community Health. 2016;70(2):168–73.

8. Mekonnen H, Medhin G, Tomlinson M, Alem A, Prince M, Hanlon C. Impact of maternal common mental disorders on child educational outcomes at 7 and 9 years: a population-based cohort study in Ethiopia. BMJ Open. 2018;8(1):e018916.

9. Rahman A, Fisher J, Bower P, Luchters S, Tran T, Yasamy MT, et al. Interventions for common perinatal mental disorders in women in low- and middle-income countries: a systematic review and meta-analysis. Bull World Health Organ. 2013;91(8):593–601I.

10. Osok J, Kigamwa P, Huang K-Y, Grote N, Kumar M. Adversities and ental health needs of pregnant adolescents in Kenya: identifying interpersonal, practical, and cultural barriers to care. BMC Women’s Health. 2018;18(1):N.PAG-N.PAG.

11. Mutua J, Kigamwa P, Ng’ang’a P, Tele A, Kumar M. A comparative study of postpartum anxiety and depression in mothers with pre-term births in Kenya. Journal of Affective Disorders Reports. 2020;2:100043.

12. Mwangi W, Gachuno O, Desai M, Obor D, Were V, Odhiambo F, et al. Uptake of skilled attendance along the continuum of care in rural Western Kenya: selected analysis from Global Health initiative survey-2012. BMC Pregnancy and Childbirth. 2018;18(1):175.

13. World Health Organization. Postnatal Care for Mothers and Newborns. Highlights from the World Health Organization 2013 Guidelines 2015.

14. Sheena Posey Norris EHF, and Bruce M. Altevogt Providing Sustainable Mental and Neurological Health Care in Ghana and Kenya: Workshop Summary. Norris SP, Forstag EH, Altevogt BM, editors. Washington, DC: The National Academies Press; 2016. 246 p.

15. Creswell J. Research Design: Qualitative, Quantitative, and Mixed-Method Approaches. 2009.

16. Odhiambo FO, Laserson KF, Sewe M, Hamel MJ, Feikin DR, Adazu K, et al. Profile: the KEMRI/CDC Health and Demographic Surveillance System--Western Kenya. Int J Epidemiol. 2012;41(4):977–87.

17. Mutea L, Ontiri S, Macharia S, Tzobotaro M, Ajema C, Odiara V, et al. Evaluating the effectiveness of a combined approach to improve utilization of adolescent sexual reproductive health services in Kenya: a quasi-experimental design study protocol. Reproductive Health. 2019;16(1):153.

18. Lavrakas PJ. Encyclopedia of Survey Research Methods SAGE 2008 1 Janurary 2011.

19. Guest G, Bunce A, Johnson L. How Many Interviews Are Enough?: An Experiment with Data Saturation and Variability. Field Methods. 2006;18(1):59–82.

20. Gale NK, Heath G, Cameron E, Rashid S, Redwood S. Using the framework method for the analysis of qualitative data in multi-disciplinary health research. BMC Medical Research Methodology. 2013;13(1):117.

21. QRS International Pty Ltd. NVivo. 2020.

22. Averill J. Matrix Analysis as a Complementary Analytic Strategy in Qualitative Inquiry. Qualitative health research. 2002;12:855–66.

23. Hambidge KM, Krebs NF. Strategies for optimizing maternal nutrition to promote infant development. Reproductive Health. 2018;15(1):87.

24. Madeghe BA, Kimani VN, Vander Stoep A, Nicodimos S, Kumar M. Postpartum depression and infant feeding practices in a low income urban settlement in Nairobi-Kenya. BMC Res Notes. 2016;9(1):506-.

25. Grote NK, Bridge JA, Gavin AR, Melville JL, Iyengar S, Katon WJ. A Meta-analysis of Depression During Pregnancy and the Risk of Preterm Birth, Low Birth Weight, and Intrauterine Growth Restriction. Archives of General Psychiatry. 2010;67(10):1012–24.

26. Stuebe AM, Grewen K, Meltzer-Brody S. Association between maternal mood and oxytocin response to breastfeeding. J Womens Health (Larchmt). 2013;22(4):352–61.

27. Nakku JEM, Nakasi G, Mirembe F. Postpartum major depression at six weeks in primary health care: prevalence and associated factors. African Health Sciences. Kampala; Uganda: Makerere University Medical School; 2006. p. 207–14.

28. Dennis C-L, McQueen K. Does maternal postpartum depressive symptomatology influence infant feeding outcomes? Acta Paediatrica. 2007;96(4):590–4.

29. Rahman A, Assad H, Rakshanda B, Siham S, Abid M, Fareed M, et al. The impact of perinatal depression on exclusive breastfeeding: a cohort study. Maternal and Child Nutrition. 2016;12(3):452–62.

30. Fathi F, Mohammad-Alizadeh-Charandabi S, Mirghafourvand M. Maternal self-efficacy, postpartum depression, and their relationship with functional status in Iranian mothers. Women & Health. 2018;58(2):188–203.

31. Mohammadi MM, Poursaberi R. The Effect of Stress Inoculation Training on Breastfeeding Self-Efficacy and Perceived Stress of Mothers With Low Birth Weight Infants: A Clinical Trial. J Family Reprod Health. 2018;12(3):160–8.

32. Jain A, Tyagi P, Kaur P, Puliyel J, Sreenivas V. Association of birth of girls with postnatal depression and exclusive breastfeeding: an observational study. BMJ open. 2014;4(6):e003545–e.

33. Annagür A, Annagür BB, Sahin A, Örs R, Kara F. Is maternal depressive symptomatology effective on success of exclusive breastfeeding during postpartum 6 weeks? Breastfeeding Medicine. 2013;8(1):53–7.

34. Figueiredo B, Canário C, Field T. Breastfeeding is negatively affected by prenatal depression and reduces postpartum depression. Psychol Med. 2014;44(5):927–36.

35. Slomian J, Honvo G, Emonts P, Reginster J-Y, Bruyère O. Consequences of maternal postpartum depression: A systematic review of maternal and infant outcomes. Womens Health (Lond). 2019;15:1745506519844044-.

36. Bell AF, Andersson E. The birth experience and women’s postnatal depression: A systematic review. Midwifery. 2016;39:112–23.

37. World Health Organization. WHO recommendations: Intrapartum care for a positive childbirth experience 2018. 200 p.

38. Vigod SN, Villegas L, Dennis CL, Ross LE. Prevalence and risk factors for postpartum depression among women with preterm and low-birth-weight infants: a systematic review. Bjog. 2010;117(5):540–50.

39. tsai AC, Tomlinson M, Comulada WS, Rotheram-Borus MJ. Food insufficiency, depression, and the modifying role of social support: Evidence from a population-based, prospective cohort of pregnant women in peri-urban South Africa. Soc Sci Med. 2016;151:69–77.

40. Rodrigues M, Patel V, Jaswal S, de Souza N. Listening to mothers: qualitative studies on motherhood and depression from Goa, India. Social science & medicine (1982). 2003;57(10):1797–806.

41. Desta M, Memiah P, Kassie B, Ketema DB, Amha H, Getaneh T, et al. Postpartum depression and its association with intimate partner violence and inadequate social support in Ethiopia: a systematic review and meta-analysis. Journal of Affective Disorders. 2021;279:737–48.

42. Were F BH, Juma D, Ayaye E, Omondi E, Boga M, Kimani M, Kiige L, Muraya KW, Murray C et al,, editor The feasibility of using peer mothers to deliver a community-based package of interventions to low birth weight infants post discharge from hospital care in Homa Bay County, Kenya. 10th KEMRI Annual Scientific and Health conference; 2020 2020; Kenya.

43. Shakya P, Kunieda MK, Koyama M, Rai SS, Miyaguchi M, Dhakal S, et al. Effectiveness of community-based peer support for mothers to improve their breastfeeding practices: A systematic review and meta-analysis. PLoS One. 2017;12(5):e0177434.

